# Artificial intelligence-based opportunistic screening for the detection of arterial hypertension through ECG signals

**DOI:** 10.1101/2022.05.14.22275082

**Authors:** Eleni Angelaki, Georgios D. Barmparis, George Kochiadakis, Spyros Maragkoudakis, Eirini Savva, Emmanuel Kampanieris, Spyros Kassotakis, Petros Kalomoirakis, Panos Vardas, Giorgos P. Tsironis, Maria E. Marketou

**Affiliations:** Institute of Theoretical and Computational Physics and Department of Physics, University of Crete, Greece; Harvard John A. Paulson School of Engineering and Applied Sciences, Harvard University, Cambridge, MA, USA; Department of Cardiology, Heraklion University Hospital, Heraklion, Greece; Department of Cardiology, Chania University Hospital, Chania, Greece; Heart Sector, Hygeia Hospitals Group, Athens, Greece

**Keywords:** electrocardiogram, hypertension, machine learning

## Abstract

**Background:** Hypertension is a major risk factor for cardiovascular disease (CVD) which often escapes the diagnosis or should be confirmed by several office visits. The electrocardiogram (ECG) is one of the most widely used diagnostic tools and could be of paramount importance in patients’ initial evaluation.

**Methods:** We used machine learning (ML) techniques based features derived from the electrocardiogram for detecting hypertension in a population without CVD. We enrolled 1091 subjects who were classified into hypertensive and normotensive group. We trained a random forest (RF), to predict the existence of hypertension in patients based only on a few basic clinical parameters and ECG-derived features. We also calculated Shapley additive explanations (SHAP), a sophisticated feature importance analysis, to interpret each feature’s role in the random forest’s predictions.

**Results:** Our RF model was able to distinguish hypertensive from normotensive patients with accuracy 84.2 %, specificity 66.7%, sensitivity 91.4%, and area under the receiver-operating curve 0.86. Age, body mass index (BMI), BMI-adjusted Cornell criteria (BMI multiplied by RaVL+SV_3_), R wave amplitude in aVL, and BMI-modified Sokolow-Lyon voltage (BMI divided by SV_1_+RV_5_), were the most important anthropometric and ECG-derived features in terms of the success of our model.

**Conclusions:** Our ML algorithm is effective in the detection of hypertension in patients using ECG-derived and basic anthropometric criteria. Our findings open new horizon in the detection of many undiagnosed hypertensive individuals who have an increased cardiovascular disease risk.

---

Hypertension is one of the most significant risk factors for cardiovascular disease (CVD) and a major cause of premature mortality and rising health care costs ^[1]^. It is a leading modifiable cause in 54% of stroke cases and 47% of ischemic heart disease incidences worldwide ^[2]^. The global prevalence of hypertensive heart disease, having risen steadily over the last decades, is expected to continue to rise due to population growth and aging ^[3]^. Unfortunately, control rates among people with hypertension are very poor, approximately 23% for women and 18% for men, with a large number of hypertensives not properly identified^[4]^.

Unawareness of hypertension is an important contributing factor to the inadequate control of the disease and absence of appropriate antihypertension treatments. Population screening programs have shown that more than 50% of hypertensives were unaware they had hypertension ^[5,6]^. Despite the progress in blood pressure (BP) measurement techniques, a substantial proportion of hypertensive patients is not identified as such, and are thus incorrectly diagnosed and managed ^[7]^. Although current practices, with the use of ambulatory and home BP measurements, have become more powerful in detecting the ‘real’ hypertensive population by discarding the white-coat effect and discovering masked hypertension, still a large proportion of patients escape diagnosis. Considering the importance of this disease for public health, exploring novel tools that potentially minimize the unawareness and increase the diagnostic performance of hypertension in daily clinical practice seems of vital importance.

The application of machine learning (ML) algorithms in the management of data is transforming the landscape of various scientific fields, including Cardiology. The fast-growing number of applications of ML/data analysis in healthcare allows identification of diseases even in early stages and prognostication of clinical outcome, thereby increasing the efficacy of treatment options. Artificial Intelligence (AI) techniques have the potential to radically change the way we practice cardiovascular medicine, providing new tools to interpret data and make aid in clinical decisions ^[8–12]^. While still a new player in Cardiology, ML has already made its mark in clinical diagnostics and research and continues to evolve rapidly ^[13–16]^. AI offers opportunities to physicians not only to make more accurate and prompt diagnoses, but also to identify hidden opportunities in improving patient management and avoiding unnecessary spending ^[17]^.

The ECG is of paramount importance in the initial evaluation of a patient suspected to have a cardiovascular pathology ^[10,12]^. In this paper we are attempting to detect whether a person is hypertensive using features from the ECG, as well as basic anthropometric features such as age, sex, and body mass index (BMI). We use digital interpretation of ECGs via computational methods and ML applications.

## METHODS

### Study population

We performed a prospective study involving two Cardiology centers from November 2019 to October 2021. We enrolled 1091 study subjects, with and without essential hypertension, and no indications of CVD. Hypertensive patients were recruited from the outpatient clinics of the respective centers. Normotensive healthy individuals were referred either for the investigation of atypical chest pain or for the modification of risk factors for cardiovascular disease such as hyperlipidemia. The diagnosis of hypertension was based on the recommendations of the European Society of Hypertension/European Society of Cardiology^[18]^; essential hypertension was defined as office BP of > 140/90 mmHg or more, measured in three consecutive visits, or in one visit when the diagnosis was confirmed by out of office measurements. In addition, out of office measurements were performed to exclude masked or white coat hypertension.

A physical examination and routine laboratory tests were performed for all subjects before inclusion. All patients underwent a routine echocardiography study. Height and weight were measured during the same visit as the ECG acquisition, and the individuals were classified using the World Health Organization (WHO) classification of BMI. We also calculated the percentage of body fat (BF) of the patients as defined by the formulae: [adult males] BF(%) = 1.20 · BMI + 0.23 · age - 10.8 - 5.4, and [adult females] BF(%) = 1.20 · BMI + 0.23 · age - 5.4 ^[19]^.

Patients with any of the following characteristics were excluded: tachy- or bradyarrhythmia; permanent atrial fibrillation, RBBB, LBBB or other conduction abnormalities on ECG, coronary artery disease; moderate or severe valvular heart disease, cardiomyopathy, cerebrovascular, liver or renal disease; history of acute coronary syndrome or myocarditis; ejection fraction < 55%; history of drug or alcohol abuse; any chronic inflammatory or other infectious disease during the last 6 months; thyroid gland disease; pregnant or lactating women. Vascular or neoplastic conditions were also ruled out vascular or neoplastic conditions were ruled out by a careful examination of the history and routine laboratory tests.

Functional tests for myocardial ischemia, coronary computed tomography angiography or invasive coronary angiography were performed according to physician’s judgement, in order to exclude coronary artery disease. The study was conducted in accordance with the Declaration of Helsinki, the protocol was approved by the Hospital Ethics Committee, and patients gave written informed consent to their participation in the study.

All numerical plots in this paper were created directly from the data, without alteration, using Python’s plotting library *matplotlib*.

### Electrocardiography

A 12-lead ECG in resting position with 10 seconds duration was performed in each subject using a digital 6-Channel machine (Biocare iE 6, Shenzhen, P.R. China) and was stored in a digital file with the eXtensive Markup Language format (XML). The sampling rate was 1000Hz. Automated measurements of wave/complex duration and wave amplitude, calculated by Biocare’s software, were extracted from the digital files. These measurements were based on representative complexes (corresponding to individual heart beats) of 1 second duration, which, according to the manufacturer, were calculated by breaking the 10 seconds signal into ten 1-second signals and averaging those into one. The final beat signals were verified, and adjusted where needed. The process was performed for each lead separately; the measurements involving time durations are averages over all leads, whereas the amplitude values are lead specific.

### ML modeling for classification

ML classification assigns a patient into two or more categories based on features used as training input to the model. Our models were trained to discriminate whether a patient is hypertensive or not, based on a number of anthropometric and ECG-derived features. We trained a logistic regression model (LR) and a *k*-nearest neighbor classifier (*k*-NN), as baselines to compare with our preferred model, the Random Forest classifier (RF).

LR estimates the (log odds) probability of a case belonging to a group/class which is expressed as a linear weighted sum of the features with which the model was trained. The *k*-NN method is a simple ML technique that does not make any statistical assumptions about the data and assigns a case to a group based on its proximity to other cases. Using cross-validation we derived that the best performing *k* was 5 (Appendix A).

A RF is a method of machine learning, an ensemble of decision trees ^[21]^. Each decision tree performs a series of binary decisions (splits) by selecting a subgroup of the input features (such as age, body fat, BMI), effectively trying out different feature order and feature combinations. A RF builds a large collection of *de-correlated* trees, and then averages their votes for the predicted class ^[22]^. RFs are good predictors even with smaller datasets due to a technique called bootstrap aggregating (bagging). Bagging trains multiple trees on overlapping, randomly selected subset of the data, and makes the final decision based on the votes of the different trees. For modeling the RF we used RandomForest classifier from scikit-learn ^[23]^. We optimized the model hyperparameters by minimizing the RF’s built-in *out-of-bag* error estimate which is almost identical to that obtained by N-fold cross-validation ^[22]^. This technique enables RFs to be trained and cross-validated in one pass. A RF is capable of handling non-linear interactions as well as correlations among features.

### Feature engineering

We calculated additional ECG waveform measurements with custom Python^[24]^ code on the 1 second representative beat produced by the electrocardiograph. Starting from the automated measurements provided by the machine, we calculated areas under curves, slopes, and heights of waveforms. Electrocardiographic terms are consistent with AMA Manual of Style (2019, 11th edition) ^[25]^.

We chose to include ECG measurements adjusted for BMI based on studies showing that larger body mass decreases the amplitude of the *R* and *S* waves in specific leads due to the electrical currents traveling longer distances. The most important ECG-derived features were: the BMI-adjusted Cornell criteria (BMI multiplied by RaVL+SV_3_);^[26]^ the BMI-modified Sokolow-Lyon voltage (BMI divided by SV_1_+RV_5_);^[12]^ and R wave amplitude in aVL.

### Feature selection

Initially we had 60 features in our dataset (Table S1). Some pairs of features exhibited high correlation, as calculated by Spearman’s rank correlation test; highly correlated features contribute the same amount of information and including both of them in a RF model might not affect performance, but it will divide, thus lessen, each feature’s predictive significance. We chose a cut-off of 0.90% correlation for removal. In our Python code we used the spearmanr function from the scipy.stats Python package. We choose the Spearman test because of the possibility of non-linear relationships among the data and then calculated the Rank-Sum statistic and ranked the features according to their p-value. This feature selection is part of pre-processing.

### Visualization

Embedding high-dimensional data into 2 dimensions allows us to visualize them in a way that gives useful insights on what differentiates study participants with a condition from those without. We performed visualization as part of our feature selection, before running any models, to inspect the features that seemed to best discriminate hypertensive from normotensive participants. A useful such method is t-distributed Stochastic Neighbor Embedding (t-SNE), a variation of Stochastic Neighbor Embedding ^[27]^ (Appendix B).

### Datasets

The dataset was randomly partitioned into a train set of 872 (80%), used directly to learn the parameters of the model, and a test set of 219 (20%), consisting of a part of the data the model had not seen before and was used exclusively for final performance evaluation of the models. Stratification for sex and history of hypertension, during the partition, ensured the two sets contained the same proportions of these two features. The ratio of hypertensive to normotensive patients in the total dataset was about 2, which did not, in our view, necessitate the use of techniques for imbalanced datasets. For validation while training the RF we used the model’s internal *out-of-bag (oob) set*. All reported performance results are on the test set. Feature importance graphs are also on the test set, as, using the train set inflates the importance of some features which might not be as important in predicting the outcome. We also made sure that data from the same patient was not included in both the train and test set.

### Feature Importance

We explored feature importance in the highest performing model which was the RF. Explaining predictions from tree models is always desired and is particularly important in medical applications, where the patterns uncovered by a model are often more important than the model’s prediction performance ^[28]^. Scikit-learn’s tree ensemble implementation allows for the computing of measures of feature importance. These measures aspire to provide insight into which features drive the model’s prediction. Mean Decrease in Impurity (MDI), an approach popular among medical researchers, calculates each feature importance as the sum over the number of splits (across all trees). It was shown that the impurity-based feature importance can inflate the role of numerical features and bias the contribution of categorical, low cardinality ones ^[29]^. Furthermore, these significances are computed on training set statistics and therefore do not reflect the usefulness of the feature in predictions that generalize to the test set. A better method is Permutation Importance which randomly shuffles a feature and calculates the error after running the model; if the error increased, then that feature is deemed important. We go one step further and calculate a recent feature importance metric called Shapley Additive explanations (SHAP), a game theoretic approach to explain the output of any machine learning model. SHAP connects optimal credit allocation with local explanations using the classic Shapley values from game theory and their related extensions ^[28,30]^. Visualizing feature importance using SHAP values is thought to be more accurate for global and local feature importance (importance calculated on *each* feature instead of all of them). SHAP values have already been used in medical papers ^[31]^.

## RESULTS

After careful screening 2156 healthy individuals, we enrolled 1091 consecutive subjects (Figure 1). Of our participants, 617 (56.55%) were female, 474 (43.45%) were male, and 712 (65.26%) were hypertensive. Overall, the mean age was 59.34*±*11.25 years old; 60.32*±*10.87 for females, and 58.07*±*11.61 for males. Based on BMI, 505 (46.29%) of them were obese, 417 (38.0%) were overweight, and 169 (15.0%) were within normal range. Compared with the normotensive group, the hypertensive group was older, had higher BMI, and tended to have slightly more female participation. The comparative statistics for a range of anthropometric and ECG features between hypertensive and normotensive population are shown in Table 1.

**TABLE 1:** Characteristics and Comparative Statistics for Hypertensive and Normotensive Study Participants.

| Feature | HTN |  |  | NT |  |  | p value |
| --- | --- | --- | --- | --- | --- | --- | --- |
|  | Mean | Std | Range | Mean | Std | Range |  |
| Age, *years* | 62.5 | 10.5 | 30.0 - 80.00 | 53.4 | 10.2 | 30.0 - 80.00 | < 0.001 |
| Body mass index, *kg/m<sup>2</sup>* | 31.4 | 5.4 | 18.8 - 56.64 | 28.1 | 5.2 | 17.6 - 48.87 | < 0.001 |
| BMI-adjusted Cornell, *mV · kg/m<sup>2</sup>* | 45.1 | 18.5 | 4.3 - 128.12 | 34.4 | 16.2 | 3.1 - 118.90 | < 0.001 |
| Body fat (1.2·BMI+0.23·age-10.8·female-5.4) | 41.6 | 9.0 | 18.9 - 75.45 | 36.6 | 8.1 | 17.3 - 67.04 | < 0.001 |
| R amplitude in aVL, *mV* | 0.6 | 0.3 | 0.0 - 1.91 | 0.5 | 0.3 | 0.0 - 1.61 | < 0.001 |
| Area under R wave in I, *ms · mV* | 9.3 | 3.9 | 0.8 - 33.36 | 7.8 | 3.3 | 0.8 - 21.10 | < 0.001 |
| QRS axis front, *degrees°* | 13.6 | 32.4 | -77.0 - 188.00 | 26.2 | 30.6 | -82.0 - 88.00 | < 0.001 |
| Corrected QT interval, *ms* | 423.7 | 24.6 | 337.0 - 500.00 | 415.0 | 22.3 | 364.0 - 506.00 | < 0.001 |
| P wave duration, *ms* | 114.6 | 15.5 | 0.0 - 196.00 | 111.7 | 10.8 | 75.0 - 149.00 | < 0.001 |
| PR interval duration, *ms* | 167.0 | 27.4 | 0.0 - 277.00 | 159.2 | 21.1 | 112.0 - 235.00 | < 0.001 |
| QT interval duration, *ms* | 397.9 | 31.7 | 290.0 - 501.00 | 387.6 | 28.8 | 312.0 - 496.00 | < 0.001 |
| R amplitude in III, *mV* | 0.2 | 0.2 | 0.0 - 1.37 | 0.3 | 0.3 | 0.0 - 1.74 | < 0.001 |
| Planar frontal QRS-T angle, *degrees°* | 37.0 | 35.3 | 0.0 - 178.00 | 26.2 | 25.3 | 0.0 - 168.00 | < 0.001 |
| Area under R wave in aVF, *ms · mV* | 3.9 | 3.6 | 0.0 - 25.39 | 4.8 | 3.7 | 0.0 - 23.51 | < 0.001 |
| Area under R wave in III, *ms · mV* | 1.9 | 2.4 | 0.0 - 21.05 | 2.7 | 3.0 | 0.0 - 19.12 | < 0.001 |
| BMI-adjusted Sokolow-Lyon voltage, *mV* | 2.6 | 0.6 | 0.2 - 5.66 | 2.4 | 0.6 | 1.0 - 4.73 | < 0.001 |
| BMI-modified Sokolow-Lyon voltage, *kg/(m<sup>2</sup> · mV)* | 17.5 | 7.7 | 4.9 - 96.88 | 15.5 | 5.9 | 5.8 - 41.94 | < 0.001 |
| Sum of QRS areas in all leads, *ms · mV* | 291.2 | 78.0 | 118.4 - 734.18 | 272.3 | 78.3 | 133.9 - 942.10 | < 0.001 |
| S amplitude in V<sub>5</sub>, *mV* | 0.4 | 0.3 | 0.0 - 1.46 | 0.3 | 0.2 | 0.0 - 1.60 | < 0.001 |
| T amplitude in V<sub>5</sub>, *mV* | 0.3 | 0.2 | -0.7 - 1.02 | 0.3 | 0.2 | -0.5 - 0.82 | < 0.001 |
| S wave amplitude in V<sub>3</sub>, *mV* | 0.9 | 0.4 | 0.0 - 2.84 | 0.8 | 0.4 | 0.0 - 3.15 | < 0.001 |
| QRS complex duration *ms* | 92.6 | 11.0 | 62.0 - 153.00 | 90.6 | 11.2 | 55.0 - 147.00 | 0.0015 |
| P amplitude in II, *mV* | 0.1 | 0.0 | 0.0 - 0.28 | 0.1 | 0.0 | 0.0 - 0.41 | 0.003 |
| J point deflection in V<sub>5</sub>, *mV* | -0.0 | 0.0 | -0.1 - 0.14 | -0.0 | 0.0 | -0.1 - 0.13 | 0.0041 |
| Q wave duration, *ms* | 10.5 | 8.1 | 0.0 - 36.00 | 11.9 | 8.6 | 0.0 - 48.00 | 0.006 |
| P axis in frontal plane, *degrees°* | 46.6 | 22.5 | -59.0 - 116.00 | 50.3 | 26.5 | -61.0 - 268.00 | 0.0065 |
| T wave duration, *ms* | 204.0 | 42.3 | 43.0 - 345.00 | 200.8 | 32.6 | 77.0 - 312.00 | 0.041 |

BMI: body mass index, HTN: participants with hypertension, NT: participants with normal blood pressure.

**FIGURE 1:**
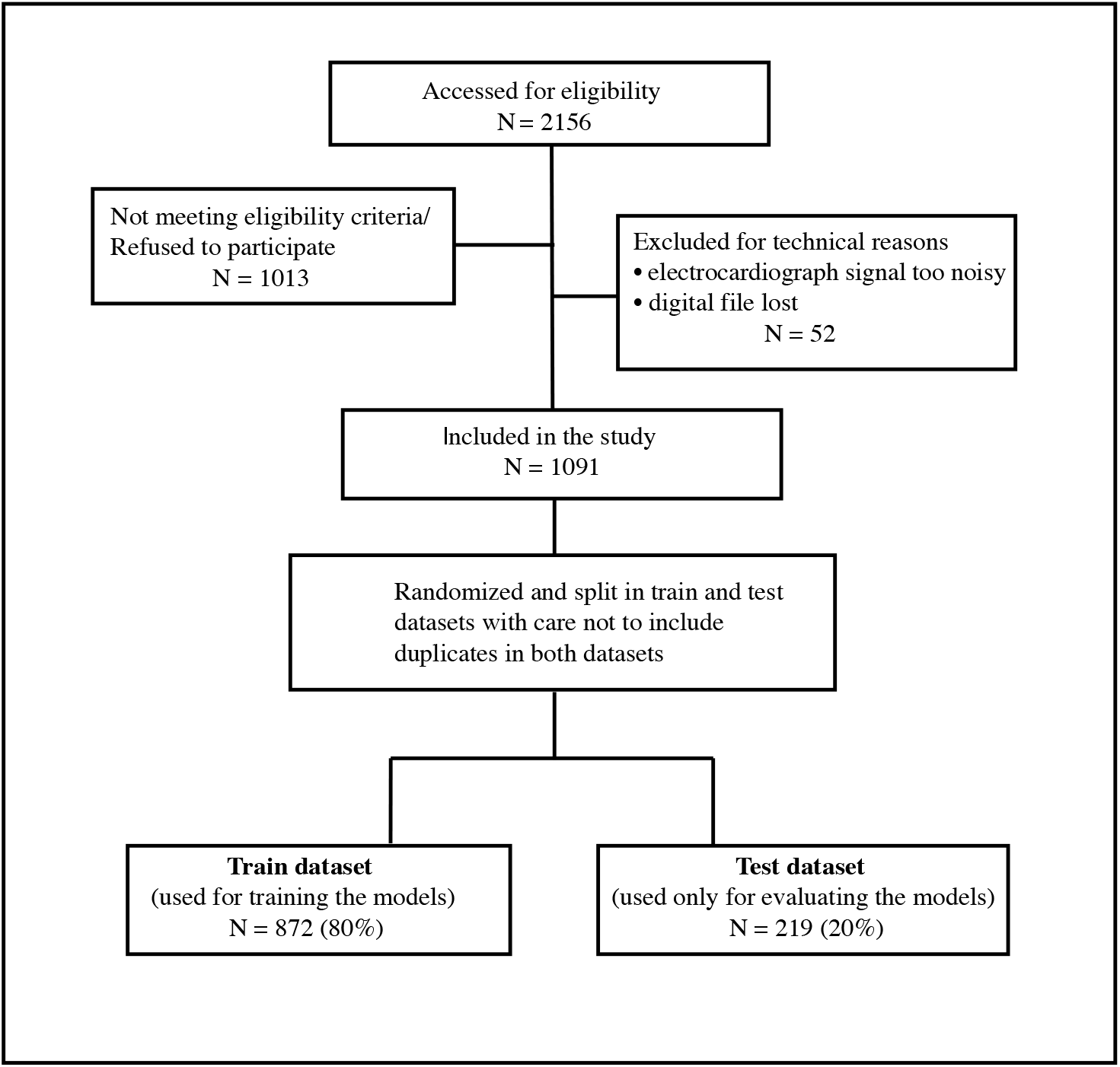
Study selection process. Study flowchart from initial hospital evaluation until data inclusion in the machine learning models.

We performed clustering with various subsets of the anthropometric and ECG features using t-SNE. In Figure 2 each point is a participant characterized by the following set of features: age, BF, BMI-adjusted Cornell criteria (BMI multiplied by RaVL+SV_3_), R wave amplitude in aVL (R_aVL), and BMI-modified Sokolow-Lyon voltage (BMI divided by SV_1_+RV_5_). This particular subset of features seems to separate hypertensive patients on the upper left corner, from normotensive patients which were in the rest of the plot.

**FIGURE 2:**
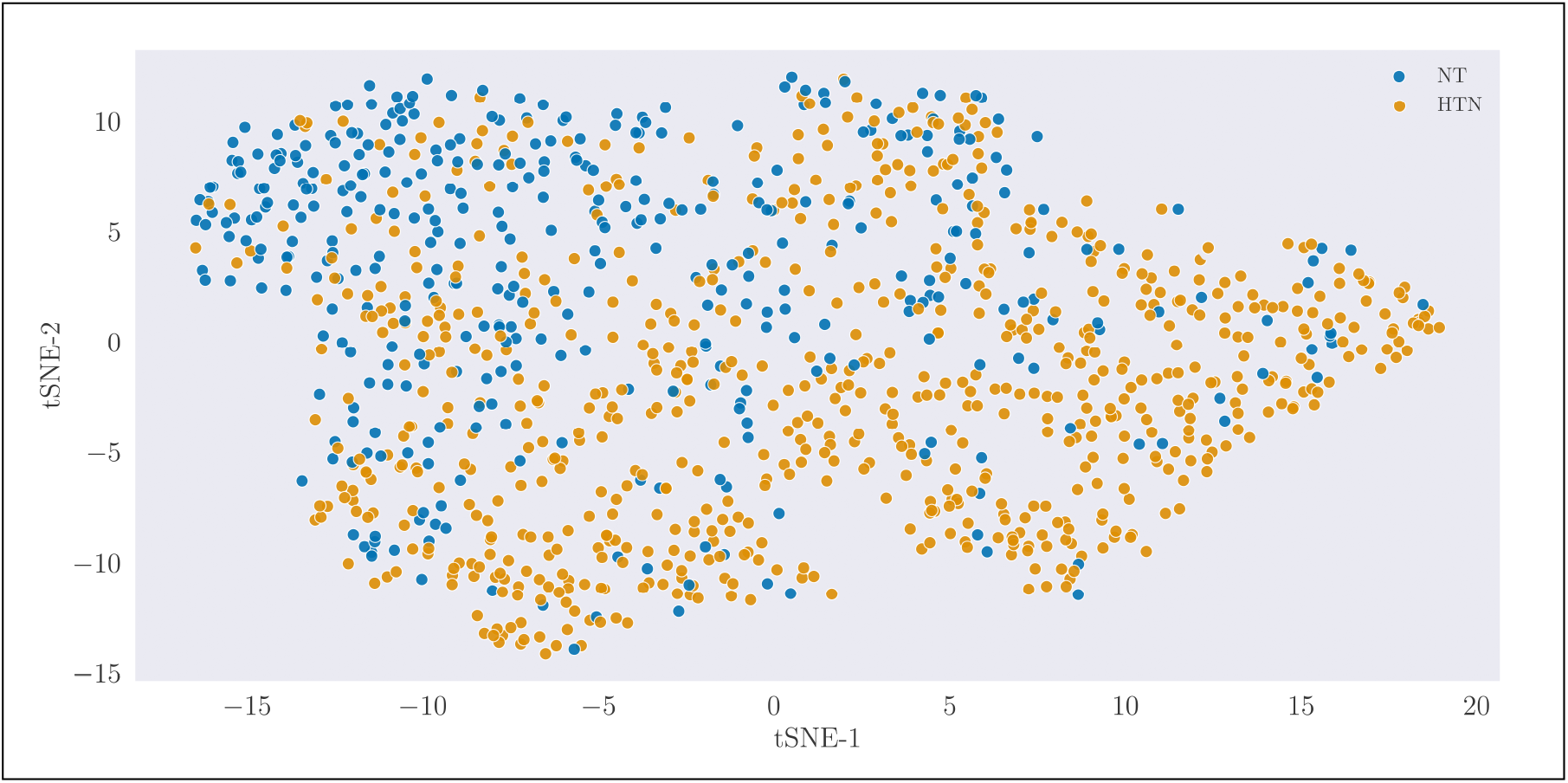
Study subject clustering using t-distributed Stochastic Neighbor Embedding (t-SNE). NT signifies the normotensive participants and HTN the hypertensive. The axes of the 2-dimensional space are given in arbitrary units.

Based on the discriminatory ability of these features, we depict the distributions of BMI, R wave amplitude in aVL, BMI-adjusted Cornell criteria (BMI multiplied by RaVL+SV_3_), and age, between normotensive and hypertensive individuals in four separate plots (Figure 3). In each plot, we separate the distributions for male and female individuals. We notice that the distributions for BMI, age, and BMI adjusted Cornell criteria, are shifted towards larger values for hypertensive than normotensive participants.

**FIGURE 3:**
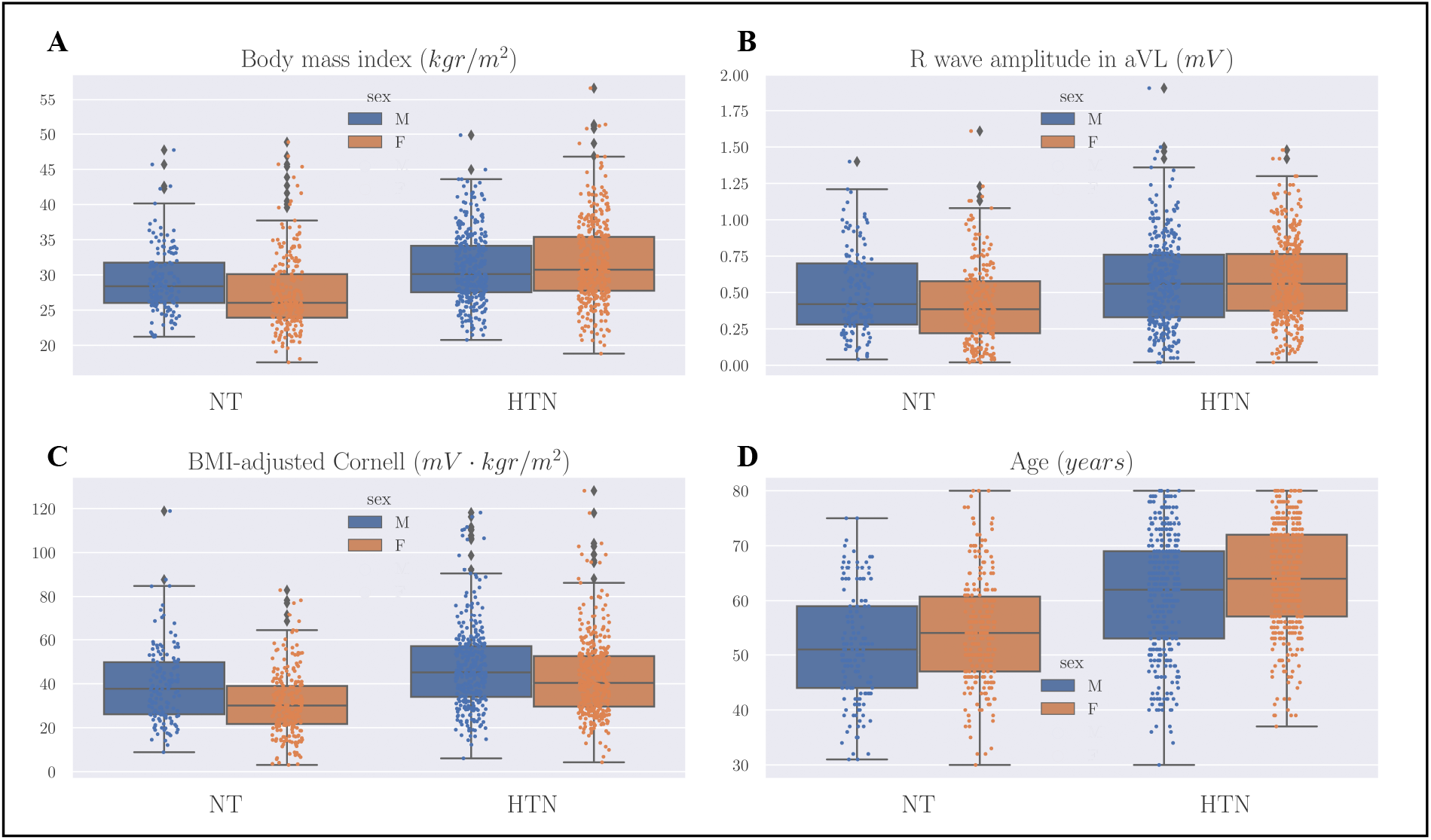
Feature distribution comparison. These box plots show the distributions of body mass index (BMI) (**A**), R wave amplitude in aVL (**B**), BMI-adjusted Cornell criteria (BMI multiplied by RaVL+SV_3_) (**C**), and age (**D**), between normotensive (NT) and hypertensive (HTN) individuals. Scatterplots (dots) of the data were superimposed for a more detailed visualization of the distributions. Each plot is also subdivided in male and female participants.

In classifying hypertensive vs. normotensive, our LR model achieved an accuracy of 77%, while the kNN classifier (*k* = 5) with 6 features, an accuracy of 78.8%. Our RF model’s accuracy was 84.2 %, specificity was equal to 66.7%, sensitivity was 91.4%, and the area under the receiver operating characteristic curve (AUC/ROC) was 0.86. All results for our models are in Table 2. Feature importance calculated by SHAP is shown in Figure 4 A. Dependence on BMI-adjusted Cornell criteria (BMI multiplied by RaVL+SV_3_) is shown in Figure 4 B. The horizontal dashed line represents the cut-off between having a negative effect on being hypertensive (below the line) and a positive one (above the line). On the x-axis we see that participants with a value above 37 *mV · kgr/m*^2^ (approximately) have a positive chance of being hypertensive. These values were calculated by SHAP on the RF model.

**TABLE 2:**
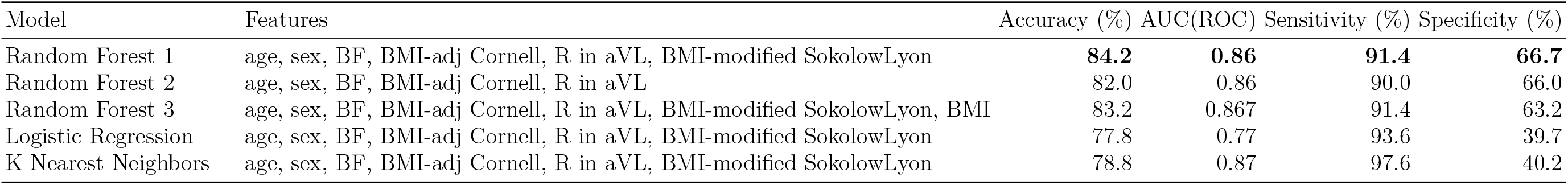
Classification performance metrics for random forest, logistic regression, and k-nearest neighbors with the respective features used in training.

**FIGURE 4:**
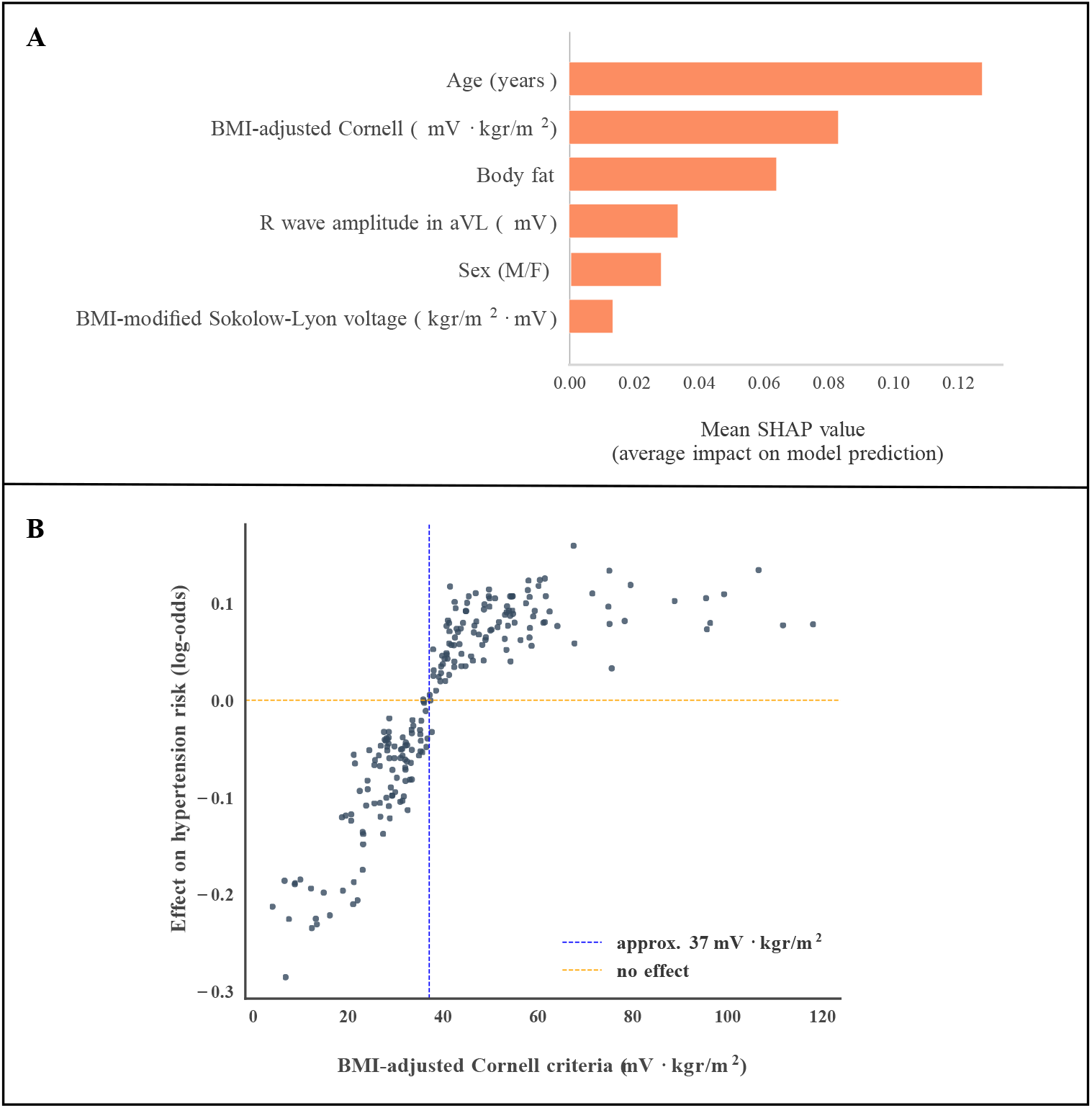
Results in detecting hypertension by the Random Forest. (**A**) Feature importances calculated on the test set using Shapley Additive explanations (SHAP). Features are body mass index (BMI), BMI-modified Sokolow-Lyon voltage (BMI divided by SV_1_+RV_5_), and BMI-adjusted Cornell criteria (BMI multiplied by RaVL+SV_3_). Body fat (BF) is defined by the formulae: [adult males] BF(%) = 1.20 BMI + 0.23 age - 10.8 - 5.4, and [adult females] BF(%) = 1.20 BMI + 0.23 age - 5.4. Sex is binary male/female (M/F). (**B**) Effect of BMI adjusted Cornell criteria on the risk of being hypertensive. Each dot in the plot is a participant whose BMI adjusted Cornell value is indicated on the *x*-axis. The values on the y-axis effectively indicate the effect of each participant’s set of features in characterizing them as hypertensive.

## DISCUSSION

To our knowledge, this is the first clinical study that exploits the promising potential of ML algorithms for the efficient and cost-effective opportunistic screening of arterial hypertension. We found specific basic clinical and ECG features that can be applied for point-of care detection of hypertensive population who will benefit from further evaluation and treatment. In our study, age, BF, BMI-adjusted Cornell criteria (BMI multiplied by RaVL+SV_3_), R wave amplitude in aVL (R_aVL), and BMI-modified Sokolow-Lyon voltage (BMI divided by SV_1_+RV_5_), seemed to separate hypertensive patients from normotensive (Figures 3 and 4A). It is remarkable that using just these features our model can detect hypertension with a good accuracy.

Our findings are very significant given that hypertension is a major public health issue ^[1]^. Although, hypertension is a leading preventable risk factor for premature death and disability worldwide, the proportion of awareness and treatment remains poor ^[5,6,18,32]^. We showed that with familiar and easily obtainable clinical tools may enhance the diagnostic efficacy and improve the detection of hypertension.

The unawareness of hypertension which is an important contributing factor for the inadequate control of the disease and often the diagnosis of hypertension is challenging and demanding even in the office. BP measurements are not always optimally performed in the routine clinical practice or even skipped altogether. Most importantly, a modest elevation in BP demands a confirmation of at least two occasions while the exclusion of clinical entities such as white coat effect or masked hypertension depends on out-of-office measurements ^[18,32]^. The increase the diagnostic performance of hypertension in daily clinical practice and the instantly derived screening at points-of care are significant for the management of the escalating burden of hypertension.

The applications of artificial intelligence on ECG are evolving rapidly with tremendous future implications on cardiovascular medicine ^[10,11,33–35]^. ECG signals and patterns largely unrecognizable to human eye interpretation can be detected by machine learning algorithms making the ECG a more powerful, non-invasive clinical tool.

Our study design has several strengths. First, the participants were carefully selected and did not have CVD since it largely eliminates other clinical parameters that could mislead our model. In this way, we improve the quality of input data and avoid various pitfalls that could arise due to the large diversity of pathological conditions that formed the basis for the training process. Second, our data were collected prospectively across real-world clinical settings and by many operators. There are limited data in the literature that attempt to associate BP level with ECG signals interpreted through ML algorithms ^[36–38]^. However, the existing knowledge has derived from limited number of ECG-data acquired from previous datasets and Physionet databases.

We present and ECG-based ML algorithm that can identify the existence of arterial hypertension by using easily obtained clinical data and ECG features in the clinical setting. This novel approach has the potential to serve as a cost-effective screening tool, empower clinicians to detect hypertensive participants and eliminate the effects of white coat and masked hypertension in the routine clinical practice. Deep learning opens up new opportunities in health care quality and advancing personalized medicine at a low cost. Our model contributes to the development of human-centered and autonomous technologies and can optimize patient-management.

Our study has some limitations. We could not control for every possible lifestyle factor, and there is possibility of residual confounding. Nonetheless, our results are clear and mainly due to the fact that our population is carefully chosen and does not have other CVD that could influence ECG features.

We did not include blood pressure levels in our analysis because the measurements were not performed at the same time with the ECG and the measurements may suffer from the white coat effect. We did not perform coronary angiogram in all participants, and this may bias the outcomes. However, we believe that this bias is small since participants underwent a meticulous work out to exclude coronary artery disease while performing coronary angiogram in low probability participants would be unethical.

The definition of hypertension was in accordance with the ESC/ESH guidelines for hypertension ^[18]^ instead of the ACC/AHA guidelines ^[39]^. This might have an impact on our results.

## PERSPECTIVES

We have showed that from basic clinical data and the use of ECG, we can identify participants with arterial hypertension. Our method offers an innovative strategy to improve health care management and personalized care at lower cost in hypertensive individuals, a population who has a high risk for CVD. ML techniques have the potential to radically change the way we practice CVD offering us novel tools to interpret data and make clinical decisions. They can also improve diagnostic performance due to the increase of the volume and complexity of the data interpreted, unlocking clinically and imaging relevant information. The capability to detect a hypertensive individual immediately and efficiently only by using a simple tool as the ECG creates great potential in the management of hypertension.

## NOVELTY AND SIGNIFICANCE

### What is New?

1. ML algorithm can efficiently identify individuals with arterial hypertension in a population without cardiovascular disease by combining clinical and ECG-extracted features.
2. BMI adjusted Cornell criteria, R wave height in aVL lead, BMI divided by the Sokolow-Lyon voltage were the most significant ECG features.

### What is Relevant?

We used machine learning (ML) techniques and found a combination of anthropometric and ECG features that enable the detection of arterial hypertension.

### Clinical/Pathophysiological Implications

ML can enhance significantly physicians’ diagnostic performance of hypertension, increase patients’ awareness and as a result improve the management of the disease.

## Supporting information

Graphical Abstract

## Data Availability

All data produced in the present study are available upon reasonable request to the authors

## Supplemental Material

Appendix A and B

Table S1

