## Supplementary material for "Artificial intelligence-based opportunistic screening for the detection of arterial hypertension through ECG signals": Graphical Abstract

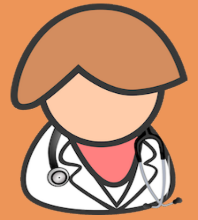

#### HYPERTENSION

A major risk factor for cardiovascular disease. Often escapes the diagnosis due to factors such as the "white coat effect" or "masked hypertension", and should be confirmed by several office visits.

#### MACHINE LEARNING: RANDOM FOREST

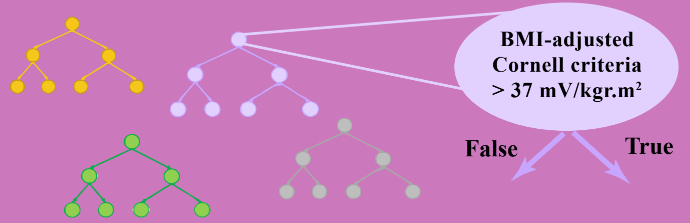


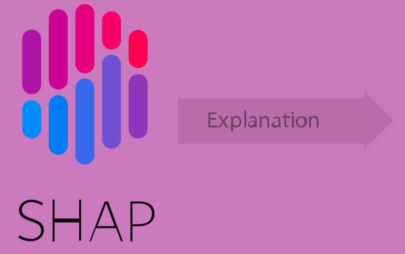


### Artificial intelligence-based opportunistic screening for the detection of arterial hypertension through ECG signals

#### Can we detect hypertension from the ECG?

**1091 Study Participants**

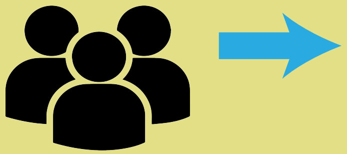

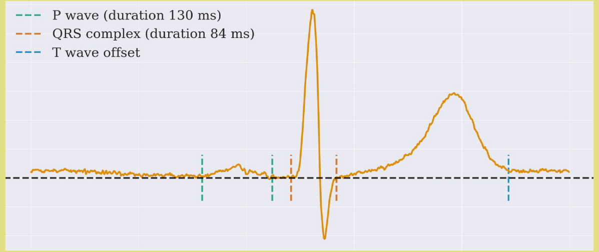


##### Feature extraction from the representative ECG waveform

BMI multiplied by RaVL+SV3
R wave amplitude in aVL
BMI divided by SV1+RV5

##### Anthropometric features

Age
Sex
Body mass index (BMI)
Body Fat

#### Feature Importance

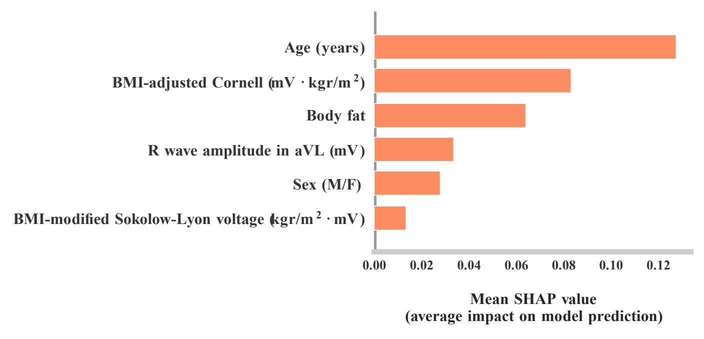


#### How does the value of BMI-adjusted Cornell criteria affect hypertension?

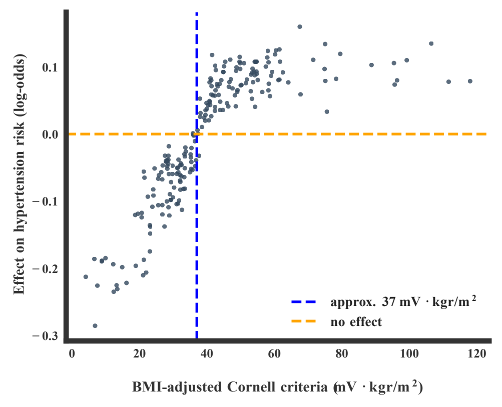


#### Patient Classification

Our machine learning algorithm was able to distinguish hypertensive from normotensive patients with accuracy 84.2 %, specificity 66.7%, sensitivity 91.4%, and area under the receiver-operating curve 0.86, using ECG-derived and very few anthropometric criteria.

*SHAP is Shapley additive explanations*
