## Supplementary material for "Artificial intelligence-based opportunistic screening for the detection of arterial hypertension through ECG signals": Supplemetary File

**Supplemental Material**

**Appendix A**

LR estimates the (log odds) probability of a case belonging to a group/class which is expressed as a linear weighted sum of the features with which the model was trained. Prior to training, features with only positive values were standardized between [0,1] using the MinMaxScaler library from the scikit-learn Python package; features with negative values, such as angles, were standardized in the interval [-1,1] using StandardScaler from the same package. For running the model we used the LogisticRegression classifier from scikit-learn with a L2 penalty term.

The $k$-NN method is used as a baseline in our case since it is a slow procedure for estimating new classifications. Every time one wants to predict the hypertensive chance of an individual, their features have to be compared with the whole dataset, in other words, one has to train the model again.

The area under the receiver operating characteristic curve (AUC/ROC) was measured using the trapezoidal rule.

**Appendix B**

Clustering method: t-distributed Stochastic Neighbor Embedding. A popular clustering method in the medical community is t-distributed Stochastic Neighbor Embedding (t-SNE), a variation of Stochastic Neighbor Embedding. The t-SNE algorithm maps points from high-dimensional space to low-dimensional space by minimizing the difference in all pairwise similarities between points in high- and low-dimensional spaces. The axes of the low-dimensional space are given in arbitrary units. The algorithm proceeds as follows: first, the pairwise distance matrix is calculated in high-dimensional space; next, the distance matrix is transformed to a similarity matrix using a varying Gaussian kernel, so that the similarity between points $X_i$ and $X_j$ represents the joint probability that $X_i$ will choose $X_j$ as its neighbor or vice versa (based on their Euclidean distance and local density); then, a random low-dimensional mapping is rendered and pairwise similarities are computed for points in the low-dimensional map. However, the low-dimensional similarities are computed using Student's t-distribution rather than a Gaussian distribution. Finally, gradient descent is used to minimize the Kullback-Leibler divergence between the two probability distributions, leading to the final low-dimensional map.

Table S 1: **Characteristics and Comparative Statistics for Hypertensive and Normotensive Study Participants.**

| Feature | HTN | | | NT | | | p value |
|---|---|---|---|---|---|---|---|
| | Mean | Std | Range | Mean | Std | Range | |
| Age, *years* | 62.5 | 10.5 | 30.0 - 80.00 | 53.4 | 10.2 | 30.0 - 80.00 | 1.5e-37 |
| Body mass index, $kgr/m^2$ | 31.4 | 5.4 | 18.8 - 56.64 | 28.1 | 5.2 | 17.6 - 48.87 | 6.8e-25 |
| BMI-adjusted Cornell, $mV \cdot kgr/m^2$ | 45.1 | 18.5 | 4.3 - 128.12 | 34.4 | 16.2 | 3.1 - 118.90 | 1.3e-22 |
| Body fat | 41.6 | 9.0 | 18.9 - 75.45 | 36.6 | 8.1 | 17.3 - 67.04 | 1.5e-18 |
| Area under R wave in aVL, $ms \cdot mV$ | 6.6 | 4.2 | 0.0 - 29.91 | 4.8 | 3.9 | 0.0 - 40.18 | 1.6e-14 |
| R wave amplitude in aVL, $mV$ | 0.6 | 0.3 | 0.0 - 1.91 | 0.5 | 0.3 | 0.0 - 1.61 | 2.4e-13 |
| Weight, $kgr$ | 86.1 | 18.0 | 43.0 - 180.00 | 78.7 | 17.4 | 45.0 - 153.00 | 5.6e-12 |
| Cornell criteria, $mV$ | 1.4 | 0.5 | 0.1 - 3.77 | 1.2 | 0.5 | 0.1 - 3.37 | 6.0e-12 |
| Area under R wave in I, $ms \cdot mV$ | 9.3 | 3.9 | 0.8 - 33.36 | 7.8 | 3.3 | 0.8 - 21.10 | 2.6e-11 |
| QRS axis front, $degrees°$ | 13.6 | 32.4 | -77.0 - 188.00 | 26.2 | 30.6 | -82.0 - 88.00 | 4.2e-11 |
| Corrected QT interval, $ms$ | 423.7 | 24.6 | 337.0 - 500.00 | 415.0 | 22.3 | 364.0 - 506.00 | 3.4e-09 |
| P wave duration, $ms$ | 114.6 | 15.5 | 0.0 - 196.00 | 111.7 | 10.8 | 75.0 - 149.00 | 6.1e-08 |
| PR interval duration, $ms$ | 167.0 | 27.4 | 0.0 - 277.00 | 159.2 | 21.1 | 112.0 - 235.00 | 8.6e-08 |
| QT interval duration, $ms$ | 397.9 | 31.7 | 290.0 - 501.00 | 387.6 | 28.8 | 312.0 - 496.00 | 9.1e-08 |
| R wave amplitude in III, $mV$ | 0.2 | 0.2 | 0.0 - 1.37 | 0.3 | 0.3 | 0.0 - 1.74 | 1.2e-07 |
| Planar frontal QRS-T angle, $degrees°$ | 37.0 | 35.3 | 0.0 - 178.00 | 26.2 | 25.3 | 0.0 - 168.00 | 3.5e-07 |
| Area under R wave in aVF, $ms \cdot mV$ | 3.9 | 3.6 | 0.0 - 25.39 | 4.8 | 3.7 | 0.0 - 23.51 | 1.2e-06 |
| Area T wave div by area of QRS complex, $ms \cdot mV$ | 1.0 | 0.5 | 0.0 - 3.46 | 1.1 | 0.5 | 0.1 - 3.12 | 1.8e-06 |
| Area under R wave in III, $ms \cdot mV$ | 1.9 | 2.4 | 0.0 - 21.05 | 2.7 | 3.0 | 0.0 - 19.12 | 2.0e-06 |
| BMI-adjusted Sokolow-Lyon voltage, $mV$ | 2.6 | 0.6 | 0.2 - 5.66 | 2.4 | 0.6 | 1.0 - 4.73 | 3.1e-06 |
| BMI-modified Sokolow-Lyon voltage, $kgr/m^2 \cdot mV$ | 17.5 | 7.7 | 4.9 - 96.88 | 15.5 | 5.9 | 5.8 - 41.94 | 3.3e-06 |
| Sum of QRS areas in all leads, $ms \cdot mV$ | 291.2 | 78.0 | 118.4 - 734.18 | 272.3 | 78.3 | 133.9 - 942.10 | 4.9e-06 |
| S wave amplitude in V_5, $mV$ | 0.4 | 0.3 | 0.0 - 1.46 | 0.3 | 0.2 | 0.0 - 1.60 | 3.7e-05 |
| T wave amplitude in V_5, $mV$ | 0.3 | 0.2 | -0.7 - 1.02 | 0.3 | 0.2 | -0.5 - 0.82 | 4.2e-05 |
| S wave amplitude in V_3, $mV$ | 0.9 | 0.4 | 0.0 - 2.84 | 0.8 | 0.4 | 0.0 - 3.15 | 2.7e-04 |
| QRS complex duration $ms$ | 92.6 | 11.0 | 62.0 - 153.00 | 90.6 | 11.2 | 55.0 - 147.00 | 1.5e-03 |
| Area under QRS interval in V_5, $ms \cdot mV$ | 34.7 | 12.3 | 10.5 - 93.14 | 32.5 | 11.1 | 11.7 - 97.14 | 2.5e-03 |
| P wave amplitude in II, $mV$ | 0.1 | 0.0 | 0.0 - 0.28 | 0.1 | 0.0 | 0.0 - 0.41 | 3.0e-03 |
| J point deflection in V_5, $mV$ | -0.0 | 0.0 | -0.1 - 0.14 | -0.0 | 0.0 | -0.1 - 0.13 | 4.1e-03 |
| Q vs. S vector | 0.9 | 0.6 | -0.8 - 2.91 | 0.9 | 0.6 | -0.8 - 2.65 | 4.2e-03 |
| Q wave duration, $ms$ | 10.5 | 8.1 | 0.0 - 36.00 | 11.9 | 8.6 | 0.0 - 48.00 | 6.0e-03 |
| P axis in frontal plane, $degrees°$ | 46.6 | 22.5 | -59.0 - 116.00 | 50.3 | 26.5 | -61.0 - 268.00 | 6.5e-03 |
| Height, $cm$ | 165.4 | 9.9 | 142.0 - 192.00 | 166.9 | 9.8 | 137.0 - 200.00 | 1.7e-02 |
| Intrincicoid deflection in II, $ms$ | 41.9 | 6.7 | 5.0 - 95.00 | 41.1 | 6.7 | 4.0 - 64.00 | 2.4e-02 |
| Area under S wave in V_1, $ms \cdot mV$ | 18.6 | 11.5 | 0.0 - 77.63 | 17.2 | 9.9 | 0.0 - 51.79 | 2.6e-02 |
| T wave amplitude in III, $mV$ | 0.0 | 0.1 | -0.4 - 0.37 | 0.0 | 0.1 | -0.3 - 0.57 | 3.6e-02 |
| T wave duration, $ms$ | 204.0 | 42.3 | 43.0 - 345.00 | 200.8 | 32.6 | 77.0 - 312.00 | 4.1e-02 |
| Heart rate, bpm | 69.9 | 11.6 | 40.0 - 129.00 | 71.1 | 12.0 | 48.0 - 109.00 | 5.4e-02 |
| R wave amplitude in V_2, $mV$ | 0.4 | 0.3 | 0.0 - 1.83 | 0.4 | 0.3 | 0.0 - 1.54 | 5.8e-02 |
| R-R interval duration, $ms$ | 884.7 | 140.5 | 464.0 - 1508.00 | 872.6 | 145.4 | 544.0 - 1360.00 | 6.7e-02 |
| T wave amplitude in aVL, $mV$ | 0.1 | 0.1 | -0.3 - 0.43 | 0.1 | 0.1 | -0.3 - 0.49 | 7.1e-02 |
| Q vs. S vector | -0.4 | 0.5 | -2.8 - 1.31 | -0.3 | 0.4 | -1.9 - 1.26 | 1.2e-01 |

BMI: body mass index, HTN: participants with hypertension, NT: participants with normal blood pressure.

(continues in next page)

(continued from previous page)

| **Feature** | **HTN** | | | **NT** | | | **p value** |
|---|---|---|---|---|---|---|---|
| | Mean | Std | Range | Mean | Std | Range | |
| ST segment duration, *ms* | 101.3 | 48.1 | 0.0 - 268.00 | 96.1 | 37.8 | 0.0 - 229.00 | 1.2e-01 |
| QRS-modified Sokolow-Lyon voltage, $ms \cdot mV$ | 187.1 | 65.3 | 26.0 - 520.02 | 182.8 | 63.3 | 79.4 - 433.00 | 1.3e-01 |
| Area under S wave in V_2, $ms \cdot mV$ | 19.5 | 12.4 | 0.0 - 89.73 | 18.3 | 11.3 | 0.0 - 72.03 | 1.3e-01 |
| P wave amplitude in V_2, *mV* | 0.0 | 0.0 | 0.0 - 0.18 | 0.0 | 0.0 | 0.0 - 0.19 | 1.4e-01 |
| T wave amplitude in V_2, *mV* | 0.3 | 0.2 | -0.3 - 1.12 | 0.3 | 0.2 | -0.2 - 1.26 | 1.9e-01 |
| S wave amplitude in V_1, *mV* | 0.7 | 0.3 | 0.0 - 2.56 | 0.7 | 0.3 | 0.0 - 1.58 | 2.1e-01 |
| Quotient of $RV_2$/RV_5 amplitudes | 0.4 | 0.3 | 0.0 - 1.99 | 0.4 | 0.3 | 0.0 - 1.71 | 2.1e-01 |
| Intrinsicoid deflection in *V*_5, *ms* | 39.6 | 6.2 | 17.0 - 103.00 | 40.0 | 6.3 | 20.0 - 66.00 | 2.2e-01 |
| R wave amplitude in V_6, *mV* | 1.1 | 0.4 | 0.0 - 2.92 | 1.1 | 0.4 | 0.0 - 3.32 | 2.4e-01 |
| Vector V_2/vector V_5 adjusted for BMI | 1.8 | 4.9 | 0.0 - 53.70 | 1.7 | 4.1 | 0.0 - 44.97 | 2.5e-01 |
| R wave amplitude in V_5, *mV* | 1.2 | 0.5 | 0.0 - 3.21 | 1.2 | 0.5 | 0.2 - 2.83 | 2.5e-01 |
| T ratio in V_5 | 1.8 | 6.1 | 0.3 - 156.00 | 1.5 | 0.6 | 0.7 - 8.00 | 2.8e-01 |
| Tallest R wave in limb leads, *mV* | 0.9 | 0.3 | 0.3 - 1.96 | 0.9 | 0.3 | 0.3 - 2.07 | 3.9e-01 |
| S wave amplitude in V_2, *mV* | 0.8 | 0.4 | 0.0 - 3.03 | 0.8 | 0.4 | 0.1 - 2.37 | 4.1e-01 |
| Area under R wave in V_5, $ms \cdot mV$ | 14.2 | 6.4 | 2.3 - 52.34 | 13.9 | 5.7 | 0.6 - 33.44 | 4.3e-01 |
| T axis frontal plane, $degrees°$ | 38.5 | 35.3 | -87.0 - 258.00 | 36.2 | 23.4 | -58.0 - 154.00 | 4.8e-01 |
| Sokolow-Lyon voltage, *mV* | 2.0 | 0.6 | 0.2 - 4.86 | 2.0 | 0.6 | 0.8 - 4.33 | 4.9e-01 |

BMI: body mass index, HTN: participants with hypertension, NT: participants with normal blood pressure.
